# Nuclear Morphometry is a superior Prognostic Predictor in comparison to Histological grading in Renal cell Carcinoma: A Retrospective Clinico-pathological study

**DOI:** 10.1101/2021.01.07.20248623

**Authors:** Shruti Agrawal, Nikunj Jain

## Abstract

**Background:** Renal cell carcinoma (RCC) comprises of a spectrum of clinico-pathologically distinct entities thereby making it difficult to accurately predict the clinical outcome. Though many predictive factors have been described in literature, tumor stage and nuclear grade have been established to consistently correlate with the tumor behaviour. However, tumors in the same stage have shown to behave differently. Similarly subjectivity and lack of reproducibility in nuclear grade mandates use of more objective parameters such as digital nuclear morphometry which could provide consistent and more reliable results in predicting prognosis. The study was conducted with the main objective of comparing the histological grade and the nuclear morphometric variables in RCC for predicting the clinical outcome.

**Material and methods:** A total of 219 cases of renal tumors in adults were retrieved retrospectively from the archives of pathology department in Sanjay Gandhi Postgraduate Institute of Medical Sciences, Lucknow and their clinical, gross and microscopic features were noted. Nuclear grading was done in 181 cases of clear cell and papillary RCC of which computer-assisted morphometry for various nuclear parameters was done in 100 cases where a follow-up data of at least 3 years was available. Nuclear grade and morphometric parameters were correlated statistically with the clinical outcome of the patients.

**Results:** Histological nuclear grade did not show statistically significant correlation with progression free survival (PFS). Higher values of mean nuclear area, mean nuclear circumference, mean nuclear major diameter and mean nuclear minor diameter were significant predictors of PFS with a strong inverse correlation.

**Conclusion:** Nuclear morphometry is a more reliable predictor of clinical outcome in patients of RCC when compared to histological grade and should be included in predictive model with other clinical and pathological parameters to accurately determine tumor behaviour.

## Introduction

Renal Cell Carcinoma (RCC) represents about 2% to 3% of all visceral cancers and accounts for 85% of the renal cancers in adults^1^. RCC is a clinicopathologically heterogenous disease which had been previously classified differently by various systems until the most recent WHO 2016 classification^2^. Despite the considerable progress made in our understanding of the pathogenesis, genetics, and pathology of RCC, predicting the clinical outcome for individual cases is challenging. The tumor stage and Fuhrman nuclear grade of the tumor have been considered as imperative factors determining the survival of these patients^3-5^. However, despite careful clinical staging, up to 30% of patients with RCC have disease progression after surgery and tumours, even at the same stage, may have different biological behaviour^6^. Similarly, subjectivity in nuclear grading and difficulty in differentiating intermediate grades contributes to the lack of uniformity in the use of nuclear grading^6,7^. Therefore, more objective methods for assessing distortion in nuclear shapes such as nuclear morphometry might prove useful for predicting prognosis for RCC. The quantitative assessment of nuclear morphometry is possible with computer imaging systems, providing a useful and a more reproducible method of predicting prognosis in renal cancer^8-10^. The purpose of the present study was to evaluate the role of nuclear morphometry in predicting disease progression-free survival, and to correlate this variable with the clinicopathological prognostic factors in RCC.

## Materials and methods

The study included consecutive cases of resected renal tumors received in the Department of Pathology from January 2014 to May 2017 where patient’s age ≥18 years. The clinical features and laboratory findings were recorded from the record maintained in hospital software system (e-hospital^@NIC^) and patient’s case files. All the cases were reviewed for gross and microscopic features. Cases of clear cell and papillary RCCs were graded according to the Fuhrman grading system into four categories^7^. Grades were revised and a secondary nuclear area was designated as ‘focal’ if it was present in <25% area and given equal weightage if it was present in 25-50% tumor area. Staging of the tumor was done according to the AJCC TNM (8^th^ edition) staging system^11^. The follow-up data of the patients was recorded based on regular OPD visits by the patients as retrieved from the hospital records as well as the data procured through contact with the patient’s family where necessary. Time of recurrence in the form of distant metastasis or local site recurrence and the time of death was noted where available.

Nuclear morphometry was assessed using a computer-assisted image-analyser system consisting of a microscope (Nikon Eclipse 80i) equipped with a high-resolution video camera (Nikon DXM1200F) and image analysis software (Image proplus, version 4.5). The highest-grade area of tumour on a representative slide from each case was selected such that they were devoid of fixation artefacts, necrotic tumour, haemorrhage and inflammation. For each patient’s sample, twenty microscopic fields were selected and 100 nuclei were digitized at a magnification of x400. The images were captured with the video camera, the nuclei viewed on the monitor, avoiding nuclei of stromal cells and the five nuclei closest to the centre of each screen outlined to minimize bias. Five morphometric descriptors were calculated for each case, i.e. the mean nuclear area (MNA), mean nuclear circumference (MNC), mean nuclear major diameter (MNMjD), mean nuclear minor diameter (MNMnD) and mean nuclear elongation factor (MNEF). Nuclear area was taken as the area enclosed inside the contour, the nuclear circumference was the contour perimeter, and the major and minor diameters were the longest and shortest orthogonal projection, respectively. Nuclear elongation factor was calculated as the ratio of major and minor diameter with a value of 1.0 for a perfect circle which increases if the nuclear contour deviates from a perfect circle. All statistical analyses were done using SPSS version 20.0 software. Correlation of qualitative factors with disease progression was done using chi-square test. Univariate analysis was done to analyse the prognostic significance of an individual factor. ROC curve analysis was used to evaluate the cut-off values and usefulness of morphological variables as a predictive marker of progression-free survival. Sensitivity, specificity and likelihood ratios were also calculated for each variable. All calculated P values were two-sided and P<0.05 was considered to indicate statistical significance.

## Results

A total of 219 consecutive cases of resected adult renal tumors between January 2014 and May 2017 were studied. These included different subtypes of RCC including clear cell carcinoma (n=154), papillary (n=27), chromophobe (n=10), mucinous tubular and spindle cell (n=3) and collecting duct carcinoma (n=1). Benign tumors comprised of oncocytoma (n=8) and papillary adenoma (n=1). Other tumors included cases of angiomyolipoma (n=7), PNET(n=3), T-cell rich B-cell lymphoma (n=1), leiomyosarcoma (n=3) and adult Wilms (n=1). The age of patients ranged from 19 to 80 years with the median age of 53.4 years and a male to female ratio of 2.7:1. Majority of the patients (58.5%, n=128) presented with abdominal pain followed by hematuria (51.6%, n=113) and abdominal lump in (36.9%, n=81). The classical triad of all the three major symptoms was present in only 8.6% (n=19) of the patients. In the laboratory investigations, haemoglobin <10g/dl was present in 73.9% cases (n=162), serum creatinine >1.5mg/dl) was present in 28.3% cases (n=62) and deranged LFT including elevated values of serum total and direct bilirubin, transaminases or alkaline phosphatase was present in 10.5% cases (n=23). Grossly, the tumor size ranged from 1.5 to 22cm with the median size of 7cm. Most tumors were primarily located in the upper pole (n=103, 47%) followed by lower pole (n=71, 32.4%) and midpole (n=16, 7.3%). Focal necrosis involving <25% of the tumor area was seen in 36.5% cases (n=80) followed by moderate (25-50%) and extensive (>50%) necrosis seen in 18.7% (n=41) and 18.3% cases (n=40) respectively.

Fuhrman nuclear grading was done based on subjective interpretation of nuclear parameters into four categories, in 181 cases including all clear cell and papillary RCCs. Thus, according to the original grades, there were 15 tumors (8.3%) with nuclear grade 1, 89 (49.2%) tumors of grade 2, 67 (37.0%) tumors of grade 3 and 10 cases (5.5%) with grade 4 nuclei including sarcomatoid differentiation. Each category was subdivided on the basis of proportion of other grades in the same tumor. According to the revised grades, there was no change in grade 1 tumors, however grade 2 tumors were reduced to 79 (43.6%), grade 3 tumors were increased to 74 (40.9%) and grade 4 tumors were increased to 13 (7.2%). Thus, nuclear grading was modified in 20 cases (Table 1). The AJCC staging was done in all cases of RCCs (n=194). Most of the tumors belonged to stage T1 (n=90, 46.4%) with size <7cm and confined to kidney while 19.6% tumors (n=38) were in stage T2 with size >7cm and confined to kidney. The second most common stage was found to be T3 (n=56, 28.9%) while only 10 cases were present in T4 stage. On grouping the TNM stages into four categories for prognostic derivations, most of the tumors constituted stage 1(n=87, 44.8%) followed by stage 3(n=58, 29.9%), stage2 (n=35, 18.0%) and stage 4 (n=14, 6.3%).

**Table 1:** Fuhrman nuclear grading of Clear cell and Papillary RCCs (n=181)

| **Fuhrman grade** | **Cases with original grade (%)** | **Subcategories** | **No. Of cases** | **Cases with revised grade (%)** |
| --- | --- | --- | --- | --- |
| **1** | 15 (8.3%) | 1 | 15 | 15 (8.3%) |
| **2** | 89 (49.2%) | 2+focal 1 | 2 | 79 (43.6%) |
|  |  | 2+1 | 10 |  |
|  |  | 2 | 67 |  |
| **3** | 67 (37.0%) | 2+focal 3 | 5 | 74 (40.9%) |
|  |  | 3+focal 2 | 7 |  |
|  |  | 3+2 | 9 |  |
|  |  | 3 | 53 |  |
| **4** | 10 (5.5%) | 3+focal 4 | 2 | 13 (7.2%) |
|  |  | 3+4 | 2 |  |
|  |  | 4 | 9 |  |

Follow-up data was available in 203 patients with a mean follow-up of 22.3 months (range 1 to 55 months). Of these, 33 patients were lost to follow-up and one was referred outside. A total of 19 patients died during the course of disease; two died in the immediate post-operative period while one died due to renal transplant-associated complications for the other kidney. Thus, follow up of >1year was available in 154 patients. Disease progression in the form of local site recurrence or distant metastasis occurred in 38 patients during a range of 11 to 20 months with a mean duration of 14.6 months and was confirmed either by imaging or re-excision surgeries. Recurrence in the form of distant metastasis occurred in lungs in 15 patients, skeletal in 5 patients, liver in 4 patients, retroperitoneal lymph nodes in 3 and bone marrow in one and brain in one patient. Local recurrence occurred in 3 patients of whom 2 presented with mass lesion in the residual kidney and one showed incisional site deposits. In the progressive group 10 patients had died, the mean duration of death from the time of surgery being 18.7 months (range 13 to 29 months) while in the non-progressive group, 6 patients had died (mean 24.6 months).

Morphometric analysis was performed in 100 cases of RCCs in which follow-up data of >1 year was available. The MNA, MNC, MNMjD and MNMnD showed significant correlation with histological type, tumour stage, nuclear grade, sarcomatoid differentiation and disease progression (Table 2). Higher values of these variables were significantly associated with sarcomatoid histology, advanced tumour stage, higher nuclear grade and tumor recurrence. In contrast, MNEF showed no significant relation to any variable except sarcomatoid differentiation. Univariate analysis showed that higher values of MNA, MNC, MNMjD and MNMnD were significant predictors of progression-free survival with a strong correlation (higher value of r). Thus, higher the values of these predictors, lesser are the chances of disease-free survival. Higher nuclear grade and tumor stage were also significant predictor of progression though they showed lesser strength of correlation. Tumor morphotype did not show significant prediction of survival in this study (Table 3). The optimal cut-off for the sensitivity and specificity values and likelihood ratios for MNA, MNC, MNMjD and MNMnD, were 150 µm^2^, 53 µm, 14 µm and 12 µm respectively. All of the variables were diagnostically useful with a likelihood ratios >1 (Table 4, Figure 1)). On Kaplan Meier survival analysis, significant difference was present in the progression-free survival (PFS) duration in patients with morphometric values more than and less than the cut-off for each of nuclear variable (Figure 2). The mean PFS was not found to be significantly different in the different nuclear grades (p=0.3) especially between grade 2 and grade 3, though mean survival appeared to be significantly different between grade 1 and grade 4. However, the revised nuclear grades into different subcategories showed a significant difference in their PFS (Figure 3, p=0.04). The stage of tumor showed significant correlation with PFS (p=0.02) however, the survival did not significantly differ in the different histological subtypes in the present study.

**Table 2:**
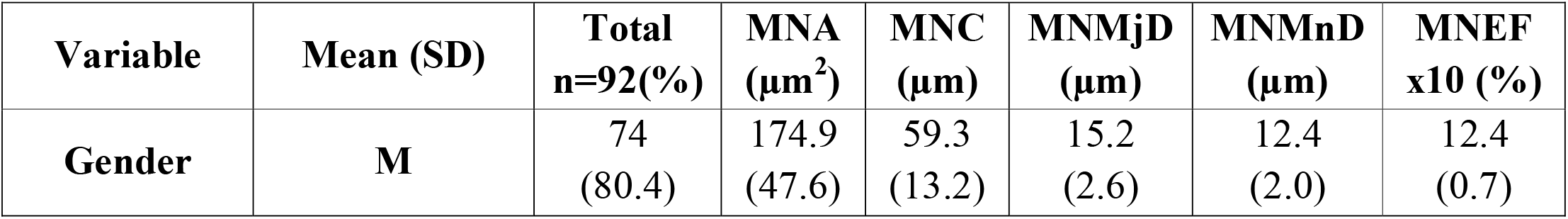
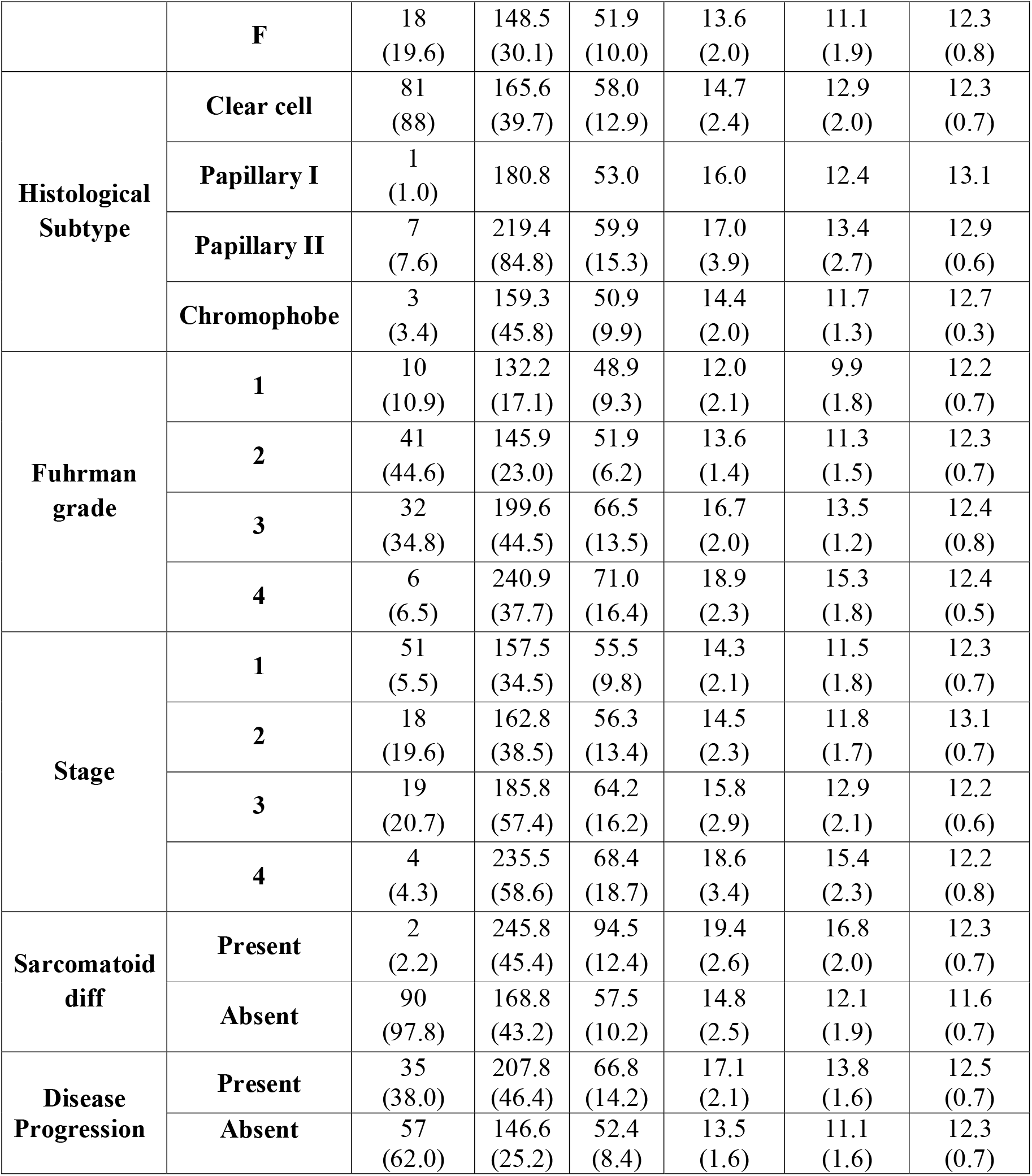
Correlation of morphometric data with histoopathological findings and disease status.

| **Variable** | **Mean (SD)** | **Total n=92(%)** | **MNA (µm<sup>2</sup>)** | **MNC (µm)** | **MNMjD (µm)** | **MNMnD (µm)** | **MNEF x10 (%)** |
| --- | --- | --- | --- | --- | --- | --- | --- |
| **Gender** | **M** | 74 (80.4) | 174.9 (47.6) | 59.3 (13.2) | 15.2 (2.6) | 12.4 (2.0) | 12.4 (0.7) |
|  | **F** | 18<br>(19.6) | 148.5<br>(30.1) | 51.9<br>(10.0) | 13.6<br>(2.0) | 11.1<br>(1.9) | 12.3<br>(0.8) |
| **Histological**<br>**Subtype** | **Clear cell** | 81<br>(88) | 165.6<br>(39.7) | 58.0<br>(12.9) | 14.7<br>(2.4) | 12.9<br>(2.0) | 12.3<br>(0.7) |
|  | **Papillary I** | 1<br>(1.0) | 180.8 | 53.0 | 16.0 | 12.4 | 13.1 |
|  | **Papillary II** | 7<br>(7.6) | 219.4<br>(84.8) | 59.9<br>(15.3) | 17.0<br>(3.9) | 13.4<br>(2.7) | 12.9<br>(0.6) |
|  | **Chromophobe** | 3<br>(3.4) | 159.3<br>(45.8) | 50.9<br>(9.9) | 14.4<br>(2.0) | 11.7<br>(1.3) | 12.7<br>(0.3) |
| **Fuhrman**<br>**grade** | **1** | 10<br>(10.9) | 132.2<br>(17.1) | 48.9<br>(9.3) | 12.0<br>(2.1) | 9.9<br>(1.8) | 12.2<br>(0.7) |
|  | **2** | 41<br>(44.6) | 145.9<br>(23.0) | 51.9<br>(6.2) | 13.6<br>(1.4) | 11.3<br>(1.5) | 12.3<br>(0.7) |
|  | **3** | 32<br>(34.8) | 199.6<br>(44.5) | 66.5<br>(13.5) | 16.7<br>(2.0) | 13.5<br>(1.2) | 12.4<br>(0.8) |
|  | **4** | 6<br>(6.5) | 240.9<br>(37.7) | 71.0<br>(16.4) | 18.9<br>(2.3) | 15.3<br>(1.8) | 12.4<br>(0.5) |
| **Stage** | **1** | 51<br>(5.5) | 157.5<br>(34.5) | 55.5<br>(9.8) | 14.3<br>(2.1) | 11.5<br>(1.8) | 12.3<br>(0.7) |
|  | **2** | 18<br>(19.6) | 162.8<br>(38.5) | 56.3<br>(13.4) | 14.5<br>(2.3) | 11.8<br>(1.7) | 13.1<br>(0.7) |
|  | **3** | 19<br>(20.7) | 185.8<br>(57.4) | 64.2<br>(16.2) | 15.8<br>(2.9) | 12.9<br>(2.1) | 12.2<br>(0.6) |
|  | **4** | 4<br>(4.3) | 235.5<br>(58.6) | 68.4<br>(18.7) | 18.6<br>(3.4) | 15.4<br>(2.3) | 12.2<br>(0.8) |
| **Sarcomatoid**<br>**diff** | **Present** | 2<br>(2.2) | 245.8<br>(45.4) | 94.5<br>(12.4) | 19.4<br>(2.6) | 16.8<br>(2.0) | 12.3<br>(0.7) |
|  | **Absent** | 90<br>(97.8) | 168.8<br>(43.2) | 57.5<br>(10.2) | 14.8<br>(2.5) | 12.1<br>(1.9) | 11.6<br>(0.7) |
| **Disease**<br>**Progression** | **Present** | 35<br>(38.0) | 207.8<br>(46.4) | 66.8<br>(14.2) | 17.1<br>(2.1) | 13.8<br>(1.6) | 12.5<br>(0.7) |
|  | **Absent** | 57<br>(62.0) | 146.6<br>(25.2) | 52.4<br>(8.4) | 13.5<br>(1.6) | 11.1<br>(1.6) | 12.3<br>(0.7) |

**Table 3:** Univariate analysis of morphometric variables and clinicopathological prognostic factors as predictors of progression.

| **Variable** | **p-value (disease-free survival)** | **r –value (strength of correlation)** |
| --- | --- | --- |
| MNA | <0.001 | -0.48 |
| MNC | <0.001 | -0.36 |
| MNMjD | <0.001 | -0.48 |
| MNMnD | <0.001 | -0.52 |
| MNEF | 0.4 | 0.07 |
| Nuclear Grade (revised) | 0.02 | -0.26 |
| Stage | 0.04 | -0.09 |
| Histological subtype | 0.2 | -0.09 |

**Table 4:** ROC analysis of morphometric variables as predictors of progression-free survival.

| **Nuclear variable** | **Cut-off value (μm)** | **Sensitivity (%)** | **Specificity (%)** | **Likelihood ratio** | **P-value** |
| --- | --- | --- | --- | --- | --- |
| **MNA** | 150 | 94.3 | 66.7 | 2.83 | <0.001 |
| **MNC** | 53 | 85.7 | 63.2 | 2.32 | <0.001 |
| **MNMjD** | 14 | 91.4 | 66.7 | 2.74 | <0.001 |
| **MNMnD** | 12 | 91.4 | 66.7 | 2.74 | <0.001 |

**Figure 1:**
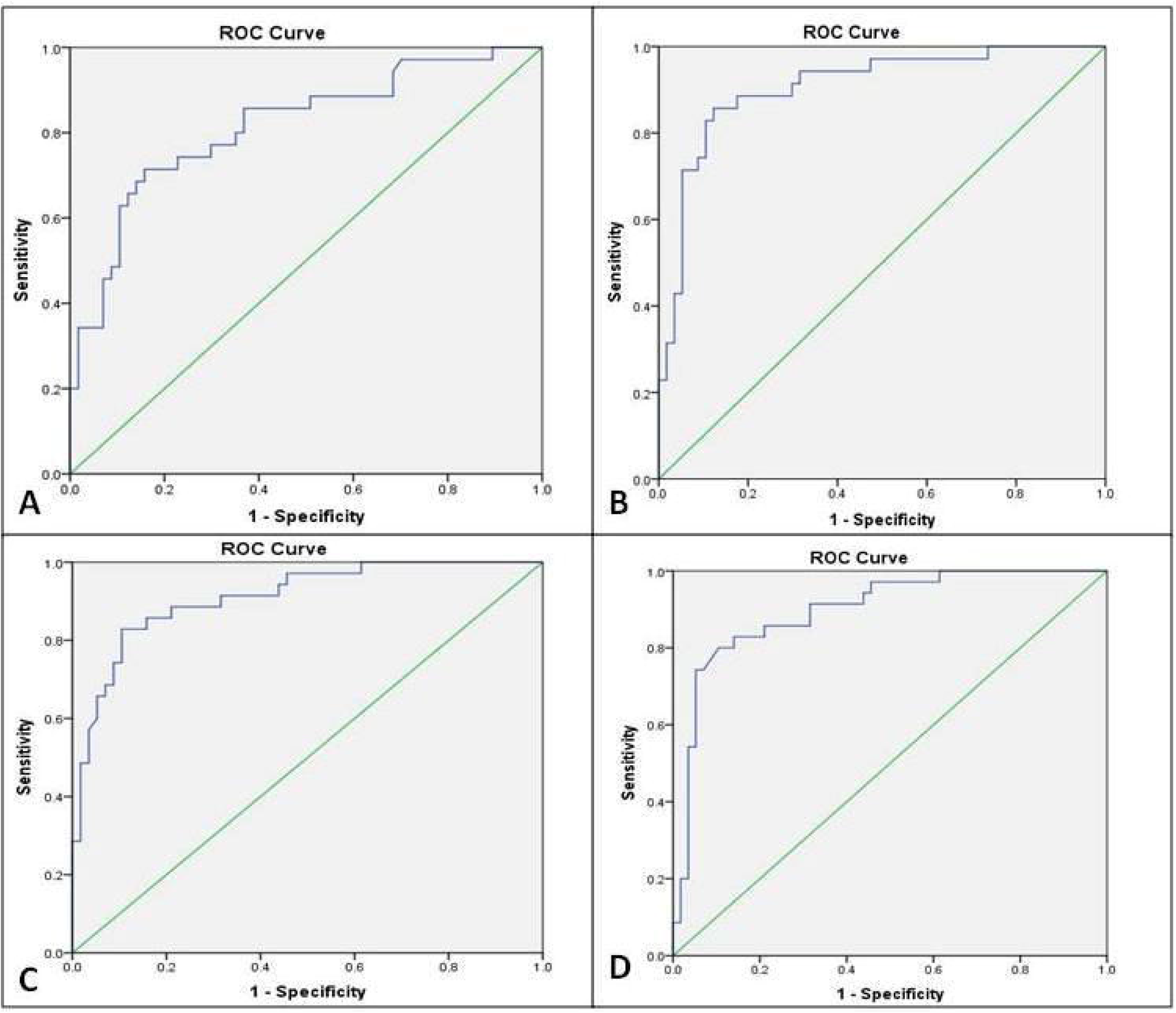
ROC curves of morphometric variables as predictors of progression-free survival. A: Mean Nuclear Area (MNA), B: Mean Nuclear circumference (MNC), C: Mean Nuclear Major Diameter(MNMjD), D: Mean Nuclear Minor Diameter (MNMnD).

**Figure 2:**
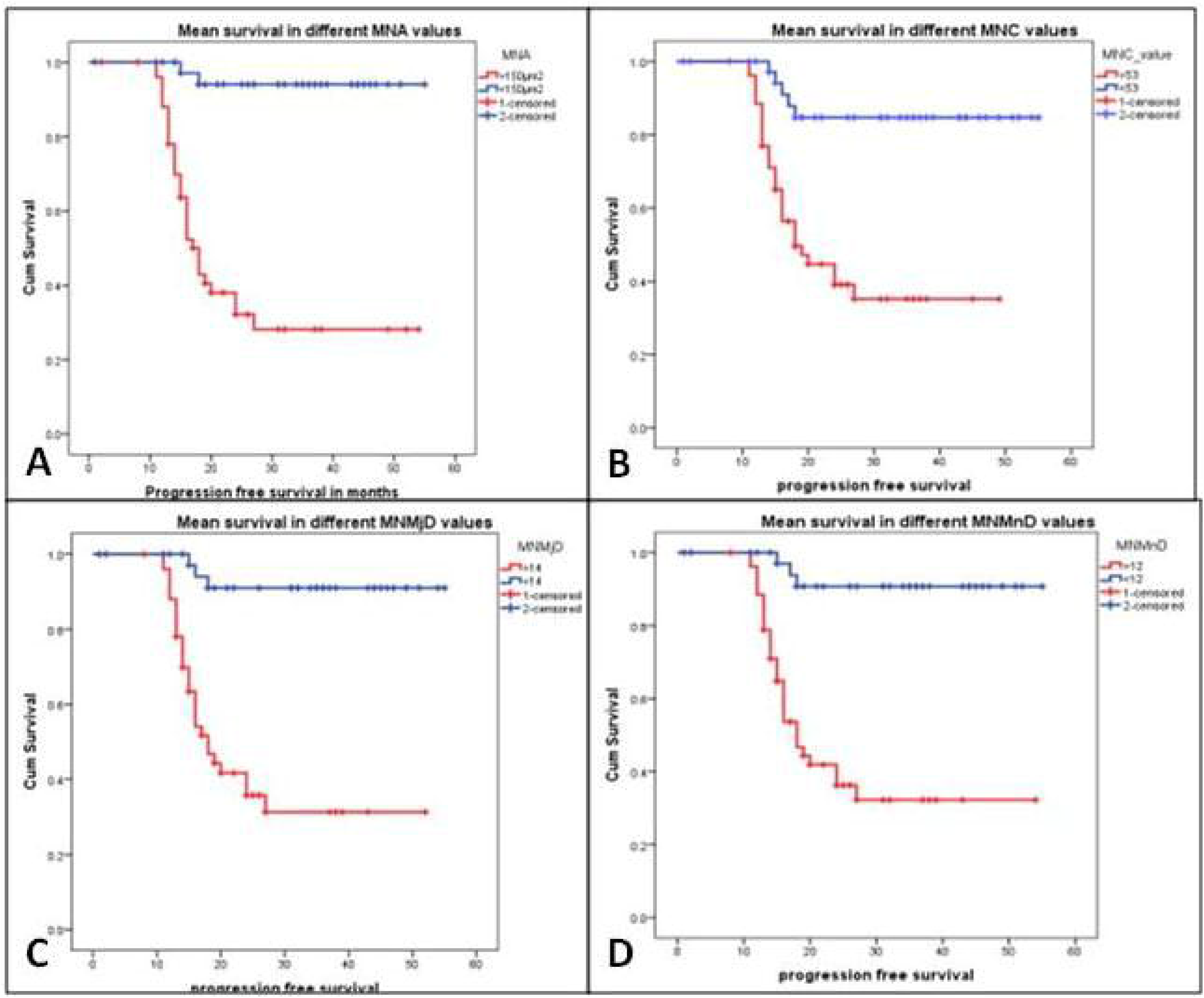
Kaplan meier plots of the probability of progression-free survival (PFS) in patients with RCC. A: Mean PFS for tumors with MNA> 150 µm^2^ vs MNA<150 µm^2^, B: Mean PFS for tumors with MNC>53 µm vs MNC, C: Mean PFS for tumors with MNMjD> 14µm vs MNMjD< 14µm, D: Mean PFS for tumors with MNMnD> 12µm vs MNMnD<12µm.

**Figure 3:**
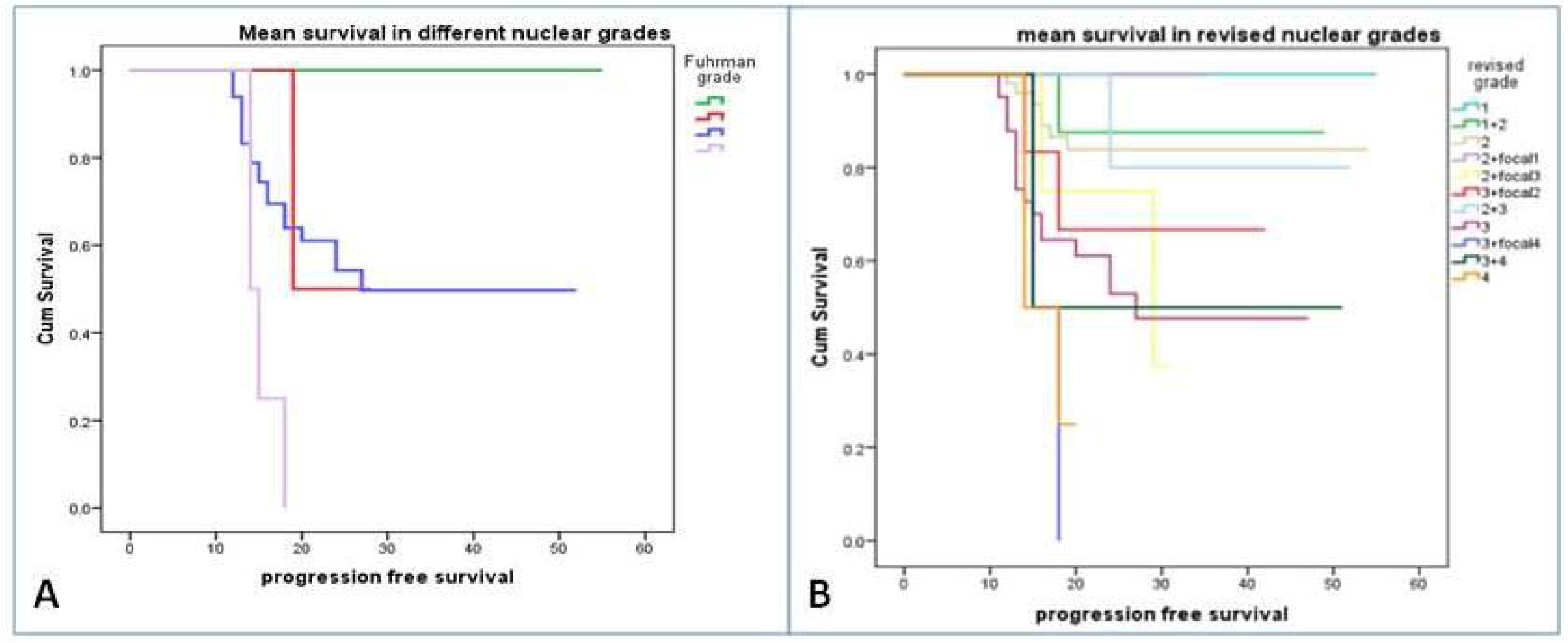
Kaplan Meier plots of the probability of PFS in different nuclear grades. A: Original nuclear grades, B: Revised nuclear grades

## Discussion

The highest incidence of RCC is found between sixth and seventh decade as reported in the literature while the mean and the median age of incidence in the present study were 53.4years and 55.0 years respectively with a range of 19 to 80 years. In the present study, the incidence of RCC was found to be more in males with a male to female ratio of 2.7:1 which was similar to the gender ratio of 2-3:1 reported in literature^12,13^. The most common presenting symptom of patients in our study was that of abdominal pain while incidentally detected cases comprised of only 2.3% as compared to 66.7% asymptomatic patients reported by Nakatani et al. and 40% incidentally detected cases in a study by sidharth et al^14,15^. The mean tumor size of 7 cm in the present study was close to 6cm and 5.3cm reported by Patard et al and Violette et al respectively^5,16^. Also, the most common location of the tumor was found to be upper pole in other studies similar to the present study^15,16^. In the histological subtype, clear cell RCC was the most frequently occurring tumor which was in concordance with the figures cited in the literature^17,18^. While microscopic tumor necrosis was identified in 73.3% of the cases in this study, it was observed in 30.5% and 40% of the tumors by sidharth et al and Stinga et al^15,18^. Though tumor necrosis has been shown to be an independent prognostic variable for RCCs, no such correlation was observed in our study^19^.

With respect to the Fuhrman nuclear grading of tumors, results in the current study were similar to other studies with most tumors displaying nuclear grade 2. However, reproducibility of the grading system was quite apparent in the study as the proportion of nuclear grades in increasing order of severity originally were 8.3%, 49.2%, 37.0 % and 5.5% respectively. After review of original grades, the revised grades showed an increase in grade 3 and grade 4 to 40.9% and 7.2% respectively while grade 2 tumors were decreased (Table 1). This highlights the subjectivity of Fuhrman grading system and imposes a limitation on the prognostic accuracy of this system as pointed in few other studies ^6,7,20^. Regarding the prognostic value of this parameter, Patard et al reported significant correlation of higher

nuclear grade (3&4) with the survival in a multivariate analysis though grade 1 and 2 lacked independent prognostic significance.^5^ Nakatani et al reported statistically significant1-year, 3-year and 5-year survival rates of 100%, 95% and 95% for grade 1 cases, 92.0%, 82.7% and 82.7% for grade 2 cases and 76.4%, 54.7% and 41.0% for grade 3 cases, respectively^14^. In the present study, Fuhrman grading did not show statistically significant difference in the progression free survival on Kaplan Meier survival analysis especially for the intermediate grades (Figure 11). On the other hand, the different categories of nuclear grade incorporating different nuclear features in the same tumor showed significant difference in the survival curves (Figure 12). This shows that including more than one kind of nuclear area in the nuclear grade enhances its accuracy of prediction though this may not be feasible if more than two cytologically different areas are present in the tumor which is not so uncommon for RCC.

Tumor staging of RCC has been established as the most consistent and powerful predictor of prognosis^4,21,22^. In the current study, most of the tumors (46.4%) presented in T1 followed by T3. This was similar to the observations of Patard et al and Violette et al^5,16^. Additionally, Patard et al found that the TNM stage group was an independent prognostic variable on multivariate analysis^5^. In the present study, the TNM stage similarly showed statistically significant prediction of survival on univariate analysis (Table 15).

Quantitative assessment of nuclear morphometry with computer imaging systems were done in various studies and correlated with the well-known prognostic factors of renal cell carcinomas^8-10,23^. According to Bektas et al, MNA, MNMjD and MNMnD and MNC moderately correlated with pathologic stages and highly correlated with Fuhrman nuclear grade. However, correlation between MNEF and Fuhrman nuclear grade, pathologic stage, and tumor size were not statistically significant^23^. In a study by Nativ et al, MNEF and MNA were the best predictors of disease free interval while all other variables including age, gender, tumor size and histological subtype did not predict disease progression rate. The 5- and 10-year survival rates were higher for patients with MNA <32µ^2^ compared with those with MNA >32µ^2^ (89% and 62% versus 50% and 40%, respectively)^8^. In the present study, the higher values of the different morphometric variables were significantly related to sarcomatoid histology, advanced tumour stage, higher nuclear grade and tumor recurrence. MNEF showed no significant relation to any variable except sarcomatoid differentiation similar to the studies done by Ozer et al and Monge et al^10,24^. Thus nuclear ellipticity does not play a significant role in deciding the aggressiveness of renal cell carcinoma. Also higher values of MNA, MNC, MNMjD and MNMnD were significant predictors of progression-free survival with a strong inverse correlation (Table 3). On Kaplan Meier survival analysis, there was significant difference in the survival of patients with morphometric values greater than cut-off value as compared to those with values lesser than the threshold (Figure 2). These findings were similar to those of Nativ et al who concluded that nuclear morphometry was prognostically superior to conventional nuclear grading in patients with localised RCC^8^.

## Conclusion

Accurate methods for identifying patients at high risk of tumour progression are clinically important. Various prognostic factors for renal cell carcinoma have been described in the literature, of which tumour stage and nuclear grade are well-established. The other factors including histological subtype, tumor necrosis and sarcomatoid morphology do not produce constant results. Though tumor stage has shown to be an independent prognostic factor in RCC, tumors even in the same stage may show different behavioural patterns. The prognostic significance of nuclear grade in separating the risk groups for recurrence is limited which is attributable partly to the subjectivity and irreproducibility of nuclear grading, particularly in the intermediate grades, and partly to the presence of different intratumoral cytological areas in a heterogenous tumor like renal cell carcinoma. This has necessitated quantitative morphometric approaches for evaluating nuclear features. Different studies including the present one prove that the higher values of different morphometric parameters are significantly related not only to the various clinicopathological factors but also to the progression-free survival of the patient. This correlation is quite expected, since these nuclear variables are the key quantitative measures of nuclear enlargement in malignant cells, and consequently included as significant criteria in most grading systems. This preliminary report confirms that a prognostic model incorporating stage and other significant factors, including nuclear morphometry, might provide significant predictive information in RCC on the clinical course, and for stratifying patients at risk in the adjuvant setting. If further prospective controlled studies validate these results, it would be reasonable to include nuclear morphometry data in future clinical research studies for patients with renal cell carcinoma.

## Data Availability

All the data mentioned in the text has been provided in the tables.

## References

1. McLaughlin JK, Lipworth L, Tarone RE. Epidemiologic aspects of renal cell carcinoma. Semin Oncol. 2006;33:527–33.

2. Moch H, Humphrey PA, Ulbright TM, Reuter VE. WHO classification of tumors of the urinary system and male genital organs.4^th^ ed. Lyon: International Agency for Research on Cancer (IARC); 2016.

3. Belldegrun AS. Renal cell carcinoma: prognostic factors and patient selection. Eur Urol. 2007;6:477–83.

4. Motzer RJ, Mazumdar M, Bacik J, Berg W, Amsterdam A, Ferrara J. Survival and prognostic stratification of 670 patients with advanced renal carcinoma. J Clin Oncol. 1999;17:2530–40.

5. Patard JJ, Leray E, Rioux-Leclercq N, Cindolo L, Ficarra V, Zisman A, De La Taille A, Tostain J, Artibani W, Abbou CC, Lobel B, Guillé F, Chopin DK, Mulders PF, Wood CG, Swanson DA, Figlin RA, Belldegrun AS, Pantuck AJ. Prognostic value of histologic subtypes in renal cell carcinoma: a multicenter experience. J Clin Oncol. 2005;23:2763–71.

6. Delahunt B. Advances and controversies in grading and staging of renal cell carcinoma. Mod Pathol. 2009;22:24–36.

7. Ficarra V, Martignoni G, Maffei N, Brunelli M, Novara G, Zanolla L, Pea M Artibani W. Original and reviewed nuclear grading according to the Fuhrman system: a multivariate analysis of 388 patients with conventional renal cell carcinoma. Cancer. 2005;103:68–75.

8. Nativ O, Sabo E, Bejar J, Halachmi S, Moskovitz B, Miselevich I. A comparison between histological grade and nuclear morphometry for predicting the clinical outcome of localized renal cell carcinoma. Br J Urol 1996; 78: 33–8.

9. Carducci MA, Piantadosi S, Pound CR, Epstein JI, Simons JW, Marshall FF, Partin AW. Nuclear morphometry adds significant prognostic information to stage and grade for renal cell carcinoma. Urology. 1999;53:44–9.

10. Ozer E, Yörükoğlu K, Sagol O, Mungan U, Demirel D, Tüzel E, Kirkali Z. Prognostic significance of nuclear morphometry in renal cell carcinoma. BJU Int. 2002;90:20–5.

11. Amin MB, Edge SB, Greene FL, et al. editors. AJCC Cancer Staging Manual. 8th ed. Switzerland: Springer, 2017.

12. Pascual D, Borque A. Epidemiology of kidney cancer. Adv Urol 2008.

13. Protzel C, Maruschke M, Hakenberg OW. Epidemiology, aetiology and pathogenesis of renal cell carcinoma. Eur Urol. 2012;11:52–9.

14. Nakatani T, Yoshida N, Iwata H, Kuratsukuri K, Uchida J, Kawashima H, Ikemoto S, Sugimura K. Clinicopathological study of renal cell carcinoma. Oncol Rep. 2003;10:679–85.

15. Sidharth, Luitel BR, Gupta DK, Maskey P, Chalise PR, Sharma UK, Gyawali PR, Shrestha GK, Sayami G, Joshi BR. Pattern of renal cell carcinoma - a single center experience in Nepal. Kathmandu Univ Med J (KUMJ). 2011 Jul-Sep;9(35):185–8.

16. Violette P, Abourbih S, Szymanski KM, Tanguay S, Aprikian A, Matthews K, Brimo F, Kassouf W. Solitary solid renal mass: can we predict malignancy? BJU Int. 2012;110:548–52

17. Karakiewicz PI, Jeldres C, Suardi N, Hutterer GC, Perrotte P, Capitanio U, Ficarra V, Cindolo L, de La Taille A, Tostain J, Mulders PF, Salomon L, Zigeuner R, Schips L, Chautard D, Valeri A, Lechevallier E, Descots JL, Lang H, Mejean A, Verhoest G, Patard JJ. Age at diagnosis is a determinant factor of renal cell carcinoma-specific survival in patients treated with nephrectomy. Can Urol Assoc J. 2008;2:610–7.

18. Stinga AC, Stinga AS, Simionescu C,Margaratescu C,Cruce M. Histopathological study of renal cell carcinoma. Curr Health Sci J. 2009;35:50–55.

19. Lee SE, Byun SS, Oh JK, et al. Significance of macroscopic tumor necrosis as a prognostic indicator for renal cell carcinoma. J Urol. 2006;176:1332–8.

20. Lang H, Lindner V, de Fromont M et al. Multicenter determination of optimal interobserver agreement using the Fuhrman grading system for renal cell carcinoma: Assessment of 241 patients with > 15-year follow-up. yCancer 2005; 103: 625-9.

21. Kim SP, Alt AL, Weight CJ, et al. Independent validation of the 2010 American Joint Committee on Cancer TNM classification for renal cell carcinoma: results from a large, single institution cohort. J Urol. 2011;185:2035–9.

22. Cheville JC, Lohse CM, Zincke H, et al. Comparisons of outcomes and prognostic feature among histological subtypes of renal cell carcinoma. Am J Surg Pathol. 2003;27:612–24.

23. Bektas S, Barut F, Kertis G, Bahadir B, Gun BD, Kandemir NO, Karadayi N, Ozdamar SO. Concordance of nuclear morphometric analysis with Fuhrman nuclear grade and pathologic stage in conventional renal cell carcinoma. Turk Patology Derg. 2008;24:14–8.

24. Monge JM, Val-Bernal JF, Buelta L, García-Castrillo L, Asensio L. Selective nuclear morphometry as a prognostic factor of survival in renal cell carcinoma. Histol Histopathol. 1999;14:119–23.

